# A nationwide Guillain-Barré syndrome epidemiological study in Spain during the COVID-19 years

**DOI:** 10.1101/2024.03.27.24304897

**Authors:** Marina Blanco-Ruiz, Lorena Martín-Aguilar, Marta Caballero-Ávila, Cinta Lleixà, Elba Pascual-Goñi, Roger Collet-Vidiella, Clara Tejada-Illa, Janina Turon-Sans, Álvaro-Carbayo, Laura Llansó, Elena Cortés, Laura Amaya Pascasio, Luis Querol

## Abstract

**OBJECTIVE:** To perform a nationwide epidemiological study of Guillain-Barré syndrome (GBS) in Spain, analysing background incidences and seasonal variation and trying to identify incidence changes during the COVID-19 years.

**METHODS:** Observational study collecting all GBS diagnoses from National Epidemiological Surveillance Network (RENAVE) collected by the Ministry of Health. Patients discharged with GBS as main diagnosis and admitted during 2018-2021 were included. Data on the incidence of SARS-CoV-2 infections and vaccinations were obtained from the National Epidemiology Centre.

**RESULTS:** In total, 3147 cases were included, 832 in 2018, 861 in 2019, 670 in 2020 and 784 in 2021. Nationwide hospital incidence was 1.78 in 2018, 1.71 in 2019, 1.41 in 2020 and 1.66 in 2021, with an increased frequency in males, elderly population, and in the winter season. Eleven percent of GBS patients needed ventilatory support. GBS and SARS-CoV-2 incidences did not correlate with one another (r=-0.29, p=0.36). GBS incidence decreased during 2020 and during COVID-19 lockdown period in comparison to the same months of 2018-2019. No relationship was found between vaccines and GBS cases during vaccination roll-out in 2021.

**INTERPRETATION:** Incidence of GBS in Spain is similar to that of other countries. Despite prior reports describing a significant increase in COVID-19-associated GBS in Spain, we detected a significant drop of GBS incidence during the SARS-CoV-2 pandemic, probably due to prevention measures. No relationship was found between SARS-CoV-2 or vaccinations and GBS incidences at the population level but data on relationship of vaccinations and GBS at the individual level were not available.

## INTRODUCTION

Guillain-Barré syndrome (GBS) is an acute, immune-mediated polyradiculopathy^1^, occurring a few days to 4 weeks after an infectious process^2^. GBS incidence ranges from 0.44 cases per 100,000 population in Japan to 2.2 cases per 100,000 population in the US^3,4^. In Spain, Cuadrado et al^5,6^ described the epidemiology of GBS in the 1990s, using data from 11 public centres distributed throughout the country, and found an incidence of 1.26 cases per 100,000 inhabitants. Other studies from smaller regions of the country described variable incidences, from 0.95 cases per 100,000 inhabitants in Cantabria^7^ to 2.07 cases per 100,000 inhabitants in Osona^8^. However, there are no studies describing nationwide GBS incidence in Spain since previous studies only covered some specific regional areas.

During the initial stages of the COVID-19 pandemic, several reports were published suggesting an association between COVID-19 and specific neurological syndromes, including GBS^9–14^. These reports mostly included small and medium size case series^15–17^. One Spanish study estimated a 5-fold increase in GBS incidence during March and April 2020 compared to the previous year, reaching 9.44 cases per 100000 people per year^18^. The authors also concluded that the relative frequency of GBS with COVID-19 was significantly higher than GBS without it. However, while it may be plausible that coronavirus infection triggers GBS, it is not clear whether the COVID-19 pandemic has led to an overall increase in GBS incidence, as it happened with other viral outbreaks such as the Zika virus pandemic^19^. In fact, a large national study of the UK population found a decrease in GBS incidence during the COVID-19 pandemic and found no association between GBS and COVID-19, although this study was performed based on the UK Immunoglobulin Database, and therefore, excluding patients treated with plasma exchange or not treated due to minor symptoms. Furthermore, one national study in Singapore showed a decreased incidence of GBS during the pandemic year, and failed to find a definitive link between GBS and COVID-19 infection^20^. On the other hand, a recent Italian study found an increase of GBS in Northern Italy in the COVID-19 era compared to the previous year^21^.

Mass SARS-CoV-2 vaccination campaigns began in 2021 raising again concerns on the possibility of an increase in GBS incidence. Several SARS-CoV-2 vaccine-associated GBS cases were reported^22–24^, especially with ChadOx1-S/nCoV-19^25^ and Ad26.COV2.S vaccines^26^ and, indeed, three national cohort studies in the UK, France and the US and one meta-analysis have found that first-dose ChAdOx1-S/nCoV-19 and Ad26.COV2.S vaccination were associated with an increased GBS risk^27–30^.

To clarify whether the COVID-19 pandemic influenced GBS incidence rates, we developed the first nationwide epidemiological study of GBS describing background GBS incidences in Spain, reporting regional and seasonal variations, and aiming to identify background GBS incidence changes during the coronavirus pandemic and mass vaccination campaigns.

## METHODS

### Study design and patients

A retrospective study extracting all patients with a primary diagnosis of GBS, included in the National Epidemiological Surveillance Network (RENAVE) of Spain (covering a population of 47 million people) during a 4-year period (2018-2021). This project has been approved by the Institutional Ethics Comittee of the Hospital de la Santa Creu i Sant Pau (IIBSP-GSB-2023-151).

The source of information was the Spanish Minimum Basic Hospital Discharge Dataset, made available by the Ministry of Health. Diagnostic and procedural coding followed the International Classification of Diseases, Tenth Revision (ICD10). Patients with a main diagnosis at discharge of GBS (ICD10 classification, codes G61.0) were included. Duplicate entries arising from hospital admissions in different institutions for the same patient were merged into a single entry. Data on the incidence of SARS-CoV-2 infection and vaccination were obtained from the database of the National Epidemiology Centre (CNE), which is part of the Ministry of Health and the Ministry of Science and Innovation.

### Statistical analysis

Continuous variables are described by mean ± SD or median [interquartile range (IQR)]. Categorical variables are described by percentages. To estimate the overall incidence of GBS, sex-specific incidence and age-specific incidence, the number of new cases each year was calculated and divided by the population at risk registered in the Spanish mid-term census, the source of the information being the National Statistics Institute.

Univariate analysis was performed with the chi-squared test or Fisher’s exact test for dichotomous variables. Continuous variables were analyzed with the t-test or the Mann– Whitney U test when appropriate. To compare incidence between years, we calculated 95% confidence intervals (CI). Simple linear regression models have been used to correlate the incidence of GBS with the incidence of SARS-CoV-2 infection and SARS-CoV-2 vaccination. To compare GBS incidences and their relationship with SARS-CoV-2 infection or vaccines we conducted a separate analysis using the months during the lockdown period in Spain (March-May) for SARS-CoV-2 and months with more vaccinations administered (June-September) for vaccines. Statistical significance for all analyses was set at 0.05 (two-sided). The analysis was carried out in SPSS 27.0 (SPSS Inc., Chicago, Illinois, USA) program for macOS. Ninety-five percent CI for incidences and proportions were performed with Stata v15.

## RESULTS

### GBS incidence

A total of 3,147 patients with GBS were detected, with the following yearly distribution: 832 in 2018, 861 in 2019, 670 in 2020 and 784 in 2021. Incidences and their 95% CI are shown in **table 1**. The mean age was 53.2 years (SD 21.9) and 63.9% of patients were male. The average incidence of GBS in Spain for the years 2018-2021 was 1.67 cases per 100,000 population. The sex-specific incidence rates were 2.18 cases per 100.000 population for males and 1.18 cases per 100.000 for females (male-to-female ratio of 2.01:1). The incidence of GBS increased with age, with the highest incidence detected at 70-79 years (3.26 [2.99-3.56]). Sex-specific and age-specific incidences rates with their 95% CI are shown in **table 1**.

**Table 1.** Incidence of GBS per year, sex-specific and age-specific incidences. n: number, CI: coefficient intervals

| Variable | Number of cases (n, %) | Incidence per 100.000 [95% CI] |
| --- | --- | --- |
| **Year** |  |  |
| 2018 | 832 (26.4%) | 1.78 [1.66-1.91] |
| 2019 | 861 (27.3%) | 1.71 [1.71-1.95] |
| 2020 | 670 (21.2%) | 1.41 [1.31-1.53] |
| 2021 | 784 (24.9%) | 1.66 [1.53-1.77] |
| **Sex** |  |  |
| Males | 2012 (63.9%) | 2.18 [2.08-2.27] |
| Females | 1135 (36.1%) | 1.18 [1.11-1.25] |
| **Age** |  |  |
| 0-9 | 168 (5.3%) | 0.97 [0.83-1.13] |
| 10-19 | 167 (5.3%) | 0.85 [0.73-0.92] |
| 20-29 | 179 (5.7%) | 0.92 [0.79-1.70] |
| 30-39 | 258 (8.2%) | 1.06[0.93-1.20] |
| 40-49 | 401 (12.7%) | 1.29 [1.16-1.42] |
| 50-59 | 542 (17.2%) | 1.93 [1.77-2.10] |
| 60-69 | 636 (20.2%) | 2.97 [2.74-3.21] |
| 70-79 | 511 (16.2%) | 3.26 [2.99-3.56] |
| 80-89 | 260 (8.3%) | 2.82 [2.48-3.18] |
| >90 | 14 (0.4%) | 0.61 [0.34-1.03] |

We also analysed the incidence in each region, with País Vasco being the region with the highest incidence (2.02 cases per 100,000 inhabitants) and La Rioja the one with the lowest incidence (1.11 cases per 100,000 inhabitants). The distribution of incidences by regions can be seen in **figure 1** and **supplementary table 1**.

**Figure 1.**
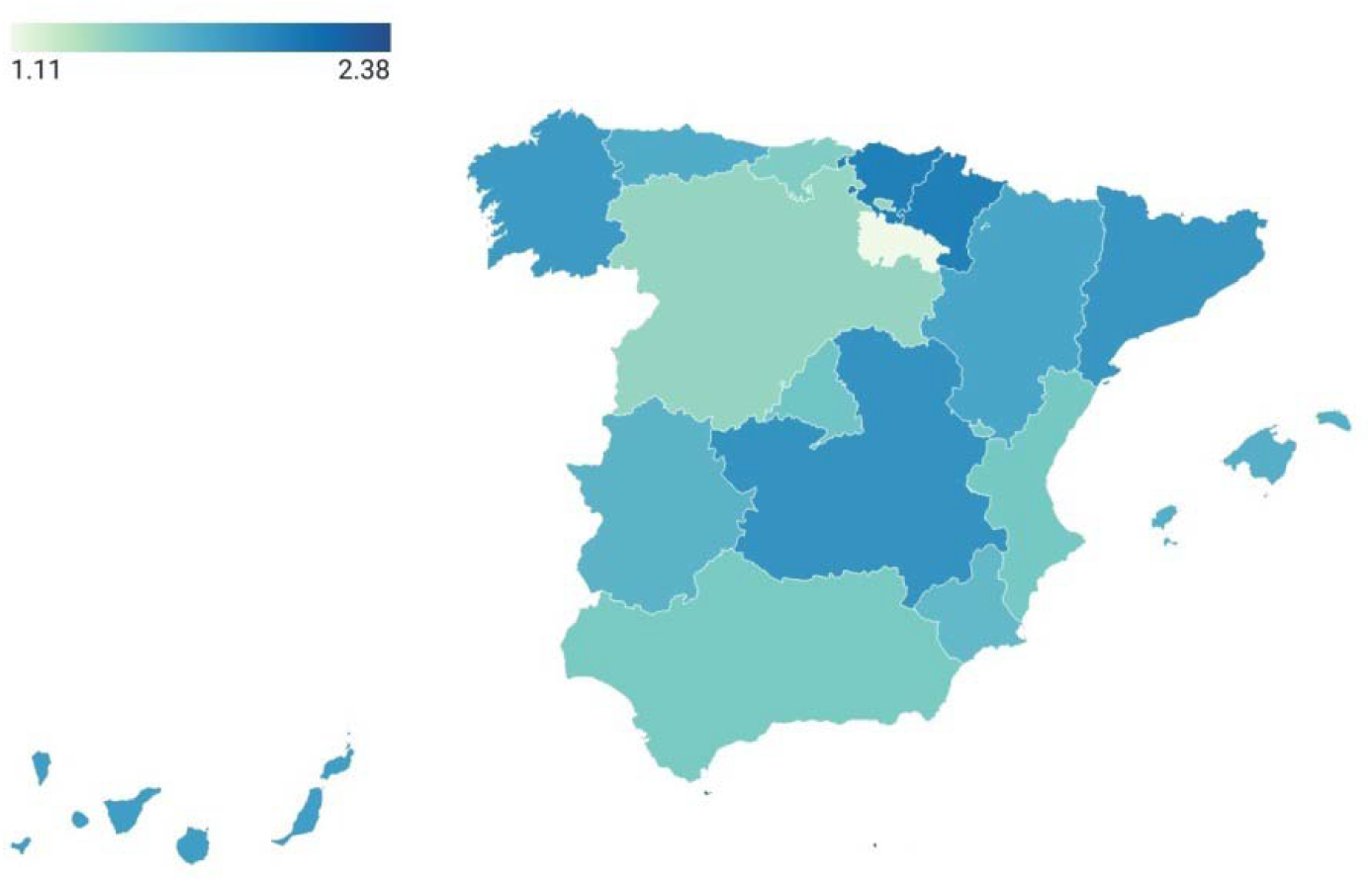
Heatmap of regional incidences of GBS.

### GBS seasonality

We found a higher frequency of GBS during winter months (**table 2**), without an overlap of their 95% CI incidences and an excess of almost 200 cases in winter (198.3). When we divide months according to the years, we can see a significant drop in GBS during the winter months of 2021, which coincides with the first winter after the pandemic (**figure 2**).

**Table 2.** Seasonal variability in GBS.

|  | Number of GBS cases | Proportion (95% CI) | p |
| --- | --- | --- | --- |
| SEASONS |  |  |  |
| Spring | 754 (24%) | 0.24 [0.22-0.25] | <0.0001 |
| Summer | 689 (21.9%) | 0.22 [0.20-0.23] |  |
| Autumm | 724 (23%) | 0.23 [0.22-0.25] |  |
| Winter | 980 (31.1%) | 0.31 [0.30-0.33] |  |
| MONTHS |  |  |  |
| January | 379 (12%) | 0.20 [0.18-0.22] | <0.0001 |
| February | 313 (9.9%) | 0.17 [0.15-0.19] |  |
| March | 289 (9.2%) | 0.15 [0.14-0.17] |  |
| April | 225 (7.1%) | 0.12 [0.10-0.14] |  |
| May | 261 (8.3%) | 0.14 [0.12-0.16] |  |
| June | 229 (7.3%) | 0.12 [0.11-0.14] |  |
| July | 227 (7.2%) | 0.12 [0.11-0.14] |  |
| August | 214 (6.8%) | 0.11 [0.10-0.13] |  |
| September | 228 (7.2%) | 0.12 [0.11-0.14] |  |
| October | 236 (7.5%) | 0.13 [0.11-0.14] |  |
| November | 241 (7.7%) | 0.13 [0.12-0.15] |  |
| December | 305 (9.7%) | 0.16 [0.14-0.18] |  |

**Figure 2.**
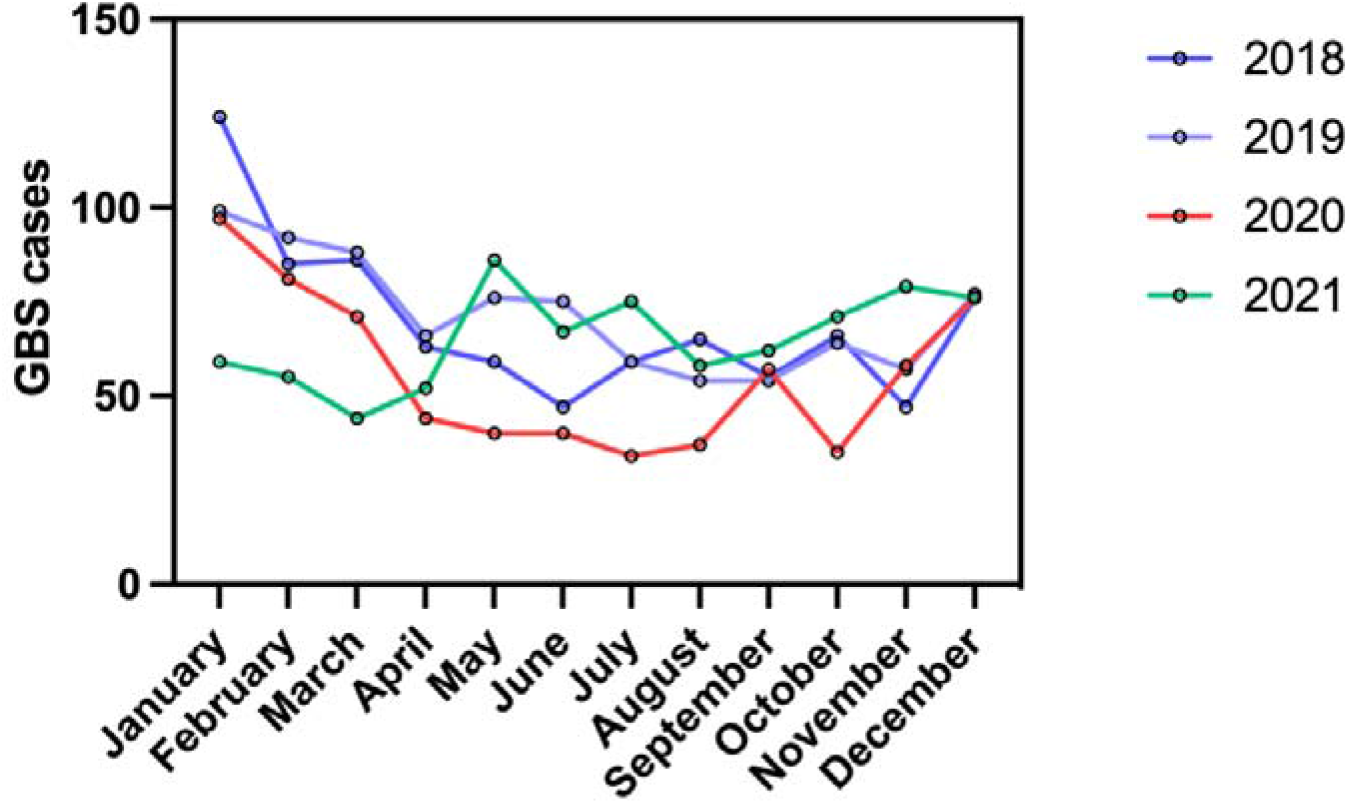
Seasonal distribution of GBS during the study years.

### GBS features

No demographic differences between GBS patients from the different study years have been found. Average hospital stay was similar between years with a median of 13 [8-23] days. Eleven percent of GBS patients needed mechanical ventilation during hospitalization and 10.4% required orotracheal intubation (OTI), with no differences between years. As for the comorbidities of the patients (**supplementary table 2**), the most frequent were hypertension (HT) (32.1%), smoking (23.1%) and diabetes (14.6%), and the most frequent infections recorded during hospitalization were respiratory, followed by urinary tract infections (UTI). Acute SARS-CoV-2 infection during hospitalization was found in less than 5% of GBS patients during 2020 and 2021 but this rate only captures concomitant and not preceding infection.

### COVID-19 and GBS

Spain was one of the European countries most affected by the COVID-19 pandemic, with one of Europe’s highest incidence and mortality rates for SARS-CoV-2, accounting for 172,541 confirmed cases and 18,056 deaths by the end of April 2020^31^. A total of 498,789 patients (1.04% of the Spanish population) were admitted to hospitals with a diagnosis of COVID-19 in the first 2 years of the pandemic.

According to CNE 1,975,439 patients in 2020 and 4,751,798 patients in 2021 had a positive PCR test for SARS-CoV-2. The lower result in 2020 is due to lower accessibility to detection tests for COVID-19, as a substantial proportion of people with symptoms compatible with COVID-19 were not tested by PCR and at least a third of infections determined by serology were asymptomatic^32^. In fact, it is estimated that during the first weeks of the pandemic in Spain, only 1 in 10 cases were detected^33^.

GBS incidence was lower during the pandemic year (1.41 [1.31-1.53] in 2020 vs 1.78 [1.66-1.91] in 2018 and 1.71 [1.71-1.95] in 2019) (**table 1**), with 116 fewer cases than expected. A decrease in the incidence was also found when comparing incidences during the lockdown period (March-May 2020) with the same months in previous years (2018-2019) (**table 3A**). Moreover, the reduction of GBS incidence was observed in all regions, including those most affected by SARS-CoV-2 at the beginning (**supplemenmtary table 3**). GBS and SARS-CoV-2 incidences did not correlate with one another (r=-0.29, p=0.36, **figure 3A**).

**Table 3.** GBS cases during SARS-CoV-2 lockdown period (A) and during maximum vaccination period (B).

| A. GBS cases during SARS-CoV-2 lockdown period (March-May) |  |  |  |  |
| --- | --- | --- | --- | --- |
| Year | Number of cases (n, %) | Study population | INC (95% CI) | p |
| 2018 | 208 (35.1%) | 46.728.814 | 0.45 [0.39-0.51] | 0.001 |
| 2019 | 230 (38.8%) | 47.105.358 | 0.49 [0.43-0.56] |  |
| 2020 | 155 (26.1%) | 47.355.684 | 0.33 [0.28-0.38] |  |
| B. GBS cases during vaccination period (June-September) |  |  |  |  |
| Year | Number of cases (n, %) | Study population | INC (95% CI) | p |
| 2018 | 226 (27%) | 46.728.814 | 0.48 [0.42-0.55] | 0.26 |
| 2019 | 242 (28%) | 47.105.358 | 0.51 [0.45-0.58] |  |
| 2021 | 262 (33%) | 47.331.302 | 0.55 [0.49-0.63] |  |

**Figure 3.**
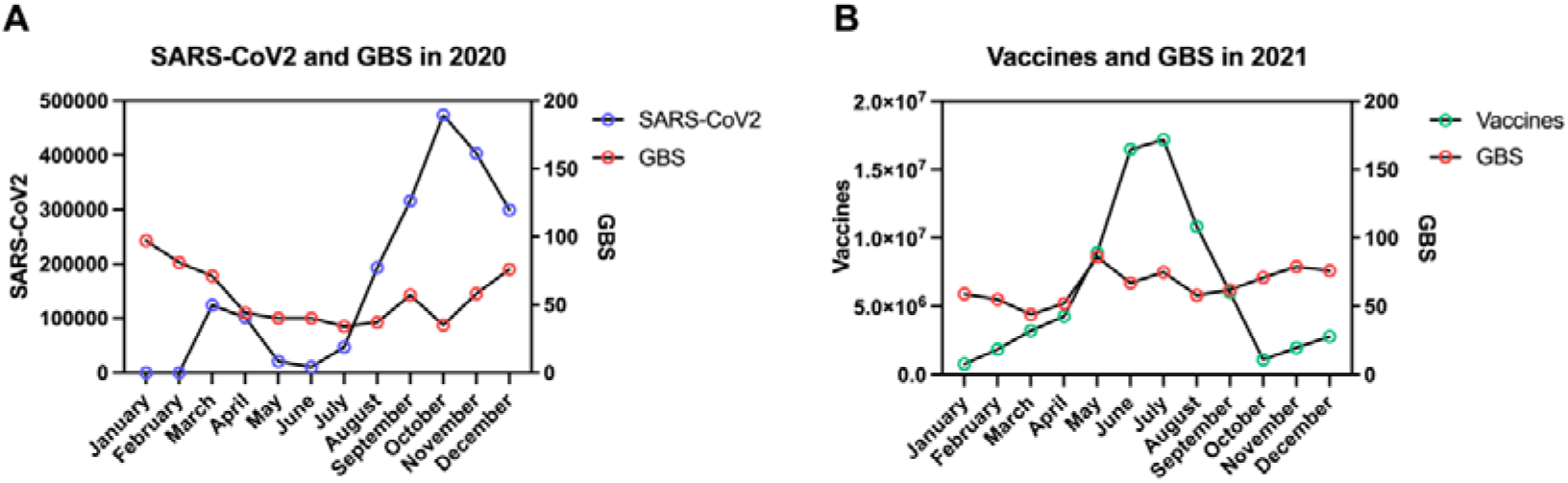
Relationship between GBS cases and SARS-CoV-2 in 2020 (A) and between GBS cases and vaccines in 2021 (B).

### Vaccines and GBS

A total of 75,512,912 vaccine doses were administered in 2021 in Spain, the most frequent being BNT162b2 (52,526,625) and mRNA-1273 (11,271,305), followed by ChAdOx1-S/nCoV-19 (9,499,075) and Ad26.COV2.S (1,977,117). To analyse the effect that mass vaccination could have had on the incidence of GBS, we selected the months with the highest number of vaccines administered (June-September) and compared data from 2021 with 2018 and 2019. We found a slight non-significant increase in GBS incidence (**table 3B**). No correlation was found between number of administered vaccines and GBS cases (**figure 3B**). Number of GBS cases and type of vaccinations are detailed in **figure 4**.

**Figure 4.**
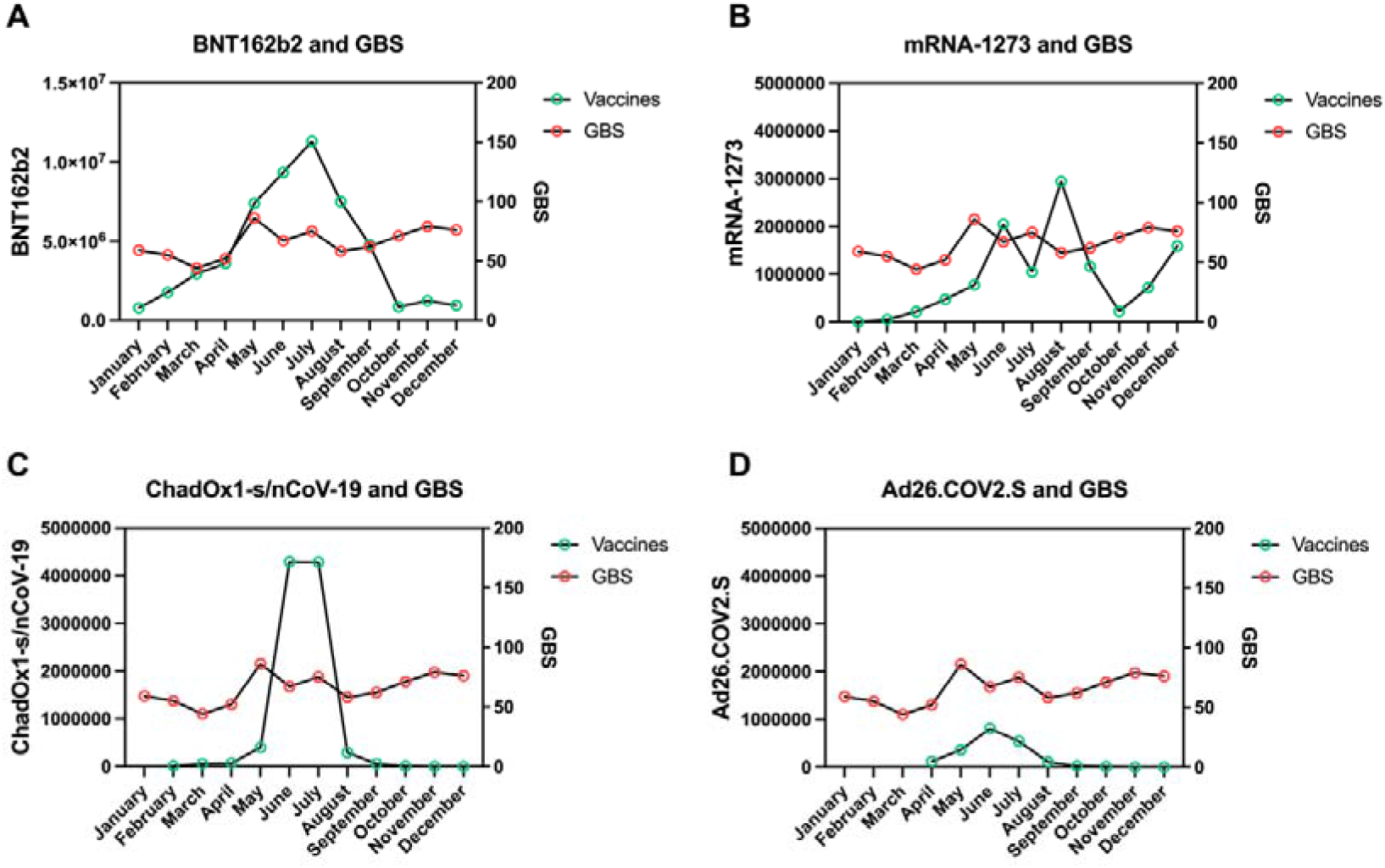
Relationship between GBS cases and different subtypes of SARS-CoV-2 vaccines: Pfizer - BNT162b2 (A), Moderna - mRNA-1273 (B), AstraZeneca - ChAdOx1-S/nCoV-19 (C) and Janssen - Ad26.COV2.S (D).

## DISCUSSION

Our study shows that GBS incidence in Spain is similar to those reported in previous studies^4^, with an increased frequency in males and in elderly population, and during winter season. GBS incidence was lower during 2020 and during the lockdown period in comparison to the same months of previous years, and we did not find an association between SARS-CoV-2 infections or vaccinations and GBS incidences at the population level.

This is the first nationwide epidemiological GBS study in Spain. We found a GBS incidence of 1.67 cases per 100,000 population, similar to previous studies in other Western countries^4^ and in Spain^5–7,34^. Our study found a significantly higher risk of GBS among males (male-to-female ratio 2:1), a finding that has been consistently demonstrated in previous studies^2,4^ and, unlike most autoimmune diseases, which usually are more frequent in women. We also found an age-dependent increase in GBS frequency, in line with previously published results describing a 20% increase every 10-year increase in age.^4^ Incidence peaked at the 70-79 age range in our study. Thus, our study confirmed that male gender and higher age are associated with GBS development^2–4^.

We also found an increased frequency in winter months, which is also consistent with prior descriptions, since typical triggering infections also follow seasonal patterns^35^. This finding has been described in Western countries, the Far East, and the Middle East, but not in the Indian subcontinent and Latin America. This lack of seasonality in some regions is partly explained by the influence of tropical climates and due to a greater predominance of a diarrhoeal prodrome or *Campylobacter jejuni* infection in low-income countries. Unfortunately, we do not have data on previous infections in our study to correlate seasonality with an antecedent respiratory infection, but only concomitant infections during hospital admission.

Our study showed no relationship between SARS-CoV-2 and GBS. There is some controversy about their relationship, as many cases and small series have been published suggesting a causal link; one national cohort study in Northern Italy reported an increase in GBS cases during the pandemic year^21^ and two national cohorts from the UK and Singapore reported a decrease^20,36^. Our study, collecting all GBS cases admitted in Spain, a country in which the pandemic hit early and that reports one of the highest COVID-19-related excess mortality rates in Western countries^37^, supports that there is no relationship between GBS and SARS-COV-2 because (1) we observe a decrease in GBS incidence during the pandemic year, probably due to prevention measures, (2) we found a decrease during lockdown period in comparison to the same months of previous years, and (3) we did not find a correlation between GBS and SARS-CoV-2 incidences. Although the GBS incidence during March-April 2020 was estimated in 9.44/100,000^18^ in a previous Spanish study from Emergency departments in COVID-19 patients, incidences remained significantly lower than the estimate (0.33/100,000) in all Spanish regions, including those with higher SARS-CoV-2 incidence. Data on GBS features were not available, but we did not find any difference in demographics, comorbidities, average hospital stay or need of ventilation in the different years studied, suggesting that the GBS pattern and the comorbidity profiles of GBS patients of the pandemic years are similar to those of non-pandemic years. Also, less than 5 percent of hospitalized GBS patients during 2020 and 2021 had a concomitant SARS-CoV-2 infection, far from the 50 percent of patients reported in the Italian cohort^21^, although information on antecedent infections was not collected in our study.

In addition, our study did not find a relationship between massive SARS-CoV-2 vaccination and GBS at the population level. However, three national cohort studies, in which individualized data link vaccinations and GBS temporally, have found an association between first-dose ChAdOx1-S/nCoV-19 and Ad26.COV2.S vaccination with an excess of GBS risk of 0.576 and 4.07 cases per 100,000 doses, respectively^27,28,30^. Also, a recently published meta-analysis showed that SARS-CoV-2 adenovirus-vectored vaccines had a 2.4-fold increased risk of GBS (seven times higher compared with mRNA-based vaccines)^29^. Individualized data, associating date of vaccination and GBS onset were not available in our study, making it difficult to find an association between them, if there is only a small excess of GBS risk. Moreover, mass vaccination in Spain mostly occurred with mRNA-based vaccines, while adenovirus-based vaccinations were significantly less common.

Several considerations and limitations should be noted in our study. First, the cohort of patients comes from the codification of diagnosis at discharge in each hospital. These diagnoses are not confirmed later on and, thus, our cohort may include some potentially misdiagnosed cases. However, it is likely that misdiagnosis rates are low in GBS, that remain stable across the years, and that the validity of our findings is not compromised by this limitation. Also, prior GBS diagnosis (unrelated to the current admission) may also have been reported in the registry. To minimize this bias, we selected only patients with GBS as the main diagnosis of the active admission, excluding all patients with prior history of GBS. Furthermore, considering we lack data on previous infections or vaccines at the individual level, we must interpret the lack of association of SARS-CoV-2 and GBS and between different types of vaccines and GBS at the description level, without inferring any lack of causality. Finally, we also lack data on clinical variants, electrophysiological subtype, or functional outcome which, with such a large cohort, would have helped defining the seasonal or yearly differences in GBS features.

In summary, this is the first nationwide epidemiological GBS study performed in Spain. Our study supports that the COVID-19 pandemic and massive SARS-CoV-2 vaccination did not increase the incidence of GBS in our country. Implementation of similar, nationwide registries, particularly if they also collect real-time and more granular, individualized information would help understand deviations in expected incidences in important diseases in the event of public health emergencies, including severe post-infectious disorders as GBS.

## Acknowledgements

We would like to thank the Registry of Specialized Health Care (RAE-CMBD) of the Ministry of Health and the Area of Health Information and Statistics of the General Subdirectorate of Health Information (MSAN). Some authors of this publication are members of the European Reference Network for rare neuromuscular diseases (EURONMD).

## Author Contributions

MB-R and LM-A analysed and interpreted the data and drafted the manuscript for intellectual content, MC-A, CL, EP-G, RC, CT, JT-S, AC, LL, EC and LA-P revised the manuscript for intellectual content. LQ designed and conceptualised the study, interpreted the data and revised the manuscript for intellectual content.

## Data Availability

Data are available upon reasonable request. All data relevant to the study are included in the article or uploaded as supplementary information. Anonymised data not published within this article will be made available by request from any qualified investigator.

## Abbreviations

CI: confidence intervals
COVID-19: coronavirus disease 2019,
GBS: Guillain-Barré syndrome
HT: hypertension
OTI: orotracheal intubation
SARS-CoV-2: severe acute respiratory syndrome coronavirus 2
SD: standard deviation,
UTI: urinary tract infection.

