## Supplementary tables for "A nationwide Guillain-Barré syndrome epidemiological study in Spain during the COVID-19 years"

| Region | 2018 Cases/ 100.000inh | 2019 Cases/ 100.000inh | 2020 Cases/ 100.000inh | 2021 Cases/ 100.000inh | Average incidence 2018-2021 |
|---|---|---|---|---|---|
| **Andalucía** | 1.50 | 1.72 | 1.28 | 1.59 | 1.52 |
| **Aragón** | 2.51 | 1.59 | 1.65 | 1.29 | 1.76 |
| **Asturias** | 2.05 | 1.57 | 1.58 | 1.69 | 1.72 |
| **Baleares** | 1.70 | 2.59 | 1.48 | 1.07 | 1.70 |
| **Canarias** | 1.96 | 1.89 | 1.56 | 1.87 | 1.82 |
| **Cantabria** | 1.72 | 1.55 | 1.55 | 1.20 | 1.50 |
| **Castilla y León** | 1.33 | 1.91 | 1.17 | 1.34 | 1.44 |
| **Castilla La Mancha** | 2.12 | 2.16 | 1.32 | 1.95 | 1.89 |
| **Cataluña** | 1.92 | 2.10 | 1.58 | 1.87 | 1.87 |
| **C. Valenciana** | 1.56 | 1.62 | 1.23 | 1.70 | 1.53 |
| **Extremadura** | 1.41 | 2.16 | 1.23 | 1.90 | 1.67 |
| **Galicia** | 2.48 | 1.70 | 1.45 | 1.71 | 1.84 |
| **Madrid** | 1.79 | 1.44 | 1.39 | 1.62 | 1.56 |
| **Murcia** | 1.49 | 2.01 | 1.39 | 1.65 | 1.63 |
| **Navarra** | 2.01 | 1.23 | 0.91 | 1.07 | 1.30 |
| **País Vasco** | 1.89 | 2.47 | 1.87 | 1.84 | 2.02 |
| **Rioja** | 0.64 | 1.91 | 0.95 | 0.95 | 1.11 |
| **Ceuta** | 1.18 | 2.37 | 4.76 | 1.21 | 2.38 |
| **Melilla** | 4.73 | 1.18 | 2.37 | 1.20 | 2.37 |
| **Total** | 1.78 | 1.83 | 1.41 | 1.66 | 1.67 |

**Supplementary table 1. Incidences of GBS per region**

| | 2018 | 2019 | 2020 | 2021 | TOTAL | p |
|---|---|---|---|---|---|---|
| **Age (mean +/- SD)** | 53.72 (21.50) | 51.74 (22.56) | 54.40 (21.55) | 52.4 (21.8) | 53.19 (21.93) | 0.15 |
| **Sex (male; n, %)** | 536 (64.4%) | 540 (62.7%) | 443 (66.1%) | 493 (62.9%) | 1519 (64.3%) | 0.49 |
| **Comorbidities (n, %)** | | | | | | |
| **HT** | 250 (30%) | 263 (30.5%) | 245 (36.6%) | 248 (31.6%) | 758 (32.1%) | 0.15 |
| **Dyslipidaemia** | 29 (3.5%) | 36 (4.2%) | 31 (4.5%) | 31 (4%) | 96 (4.1%) | 0.49 |
| **Diabetes** | 122 (14.7%) | 125 (14.5%) | 102 (15.2%) | 114 (14.5%) | 349 (14.6%) | 0.88 |
| **Smoking** | 199 (23.9%) | 205 (23.8%) | 143 (21.3%) | 197 (25.1%) | 547 (23.1%) | 0.86 |
| **Concomitant infections (n, %)** | | | | | | |
| **UTI** | 54 (6.5%) | 51 (5.9%) | 57 (8.5%) | 91 (11.6%) | 162 (6.9%) | 0.08 |
| **Respiratory infection** | 66 (7.9%) | 64 (7.4%) | 63 (9.4%) | 43 (5.5%) | 193 (8.2%) | 0.13 |
| **COVID-19** | 0 (0%) | 0 (0%) | 25 (3.7%) | 34 (4.3%) | 25 (1.1%) | <0.001 |
| **AGE** | 18 (2.2%) | 19 (2.2%) | 16 (2.4%) | 23 (2.9%) | 53 (2.2%) | 0.57 |
| **Average hospital stay (days; median, IQR)** | 12 [7-23] | 13 [7-23] | 13 [8-24] | 12 [7-25] | 13 [8-23] | 0.74 |
| **Procedures (n, %)** | | | | | | |
| **Ventilation** | 78 (9.4%) | 102 (11.8%) | 74 (11%) | 93 (11.9%) | 347 (11%) | 0.33 |
| **OTI** | 69 (8.3%) | 95 (11%) | 71 (10.6%) | 93 (11.9%) | 328 (10.4%) | 0.11 |

**Supplementary table 2. Demographic characteristics, comorbidities, and need of ventilation in GBS patients.** AGE: acute gastroenteritis; HT: hypertension; IQR: interquartile range; OTI: orotracheal intubation; SD: standard deviation; UTI: urinary tract infection.

| | 2018 | 2019 | 2020 |
|---|---|---|---|
| Andalucía | 38 | 52 | 19 |
| Aragón | 6 | 7 | 4 |
| Asturias | 3 | 6 | 7 |
| Baleares | 3 | 7 | 3 |
| Canarias | 14 | 10 | 8 |
| Cantabria | 1 | 2 | 1 |
| Castilla y León | 5 | 9 | 6 |
| Castilla la Mancha | 9 | 12 | 10 |
| Cataluña | 38 | 37 | 31 |
| Comunidad Valenciana | 17 | 22 | 17 |
| Estremadura | 5 | 2 | 2 |
| Galicia | 17 | 10 | 8 |
| Madrid | 27 | 22 | 23 |
| Murcia | 7 | 7 | 5 |
| Navarra | 5 | 5 | 1 |
| Pais Vasco | 11 | 17 | 8 |
| La Rioja | 1 | 3 | 1 |
| Melilla/Ceuta | 1 | 0 | 1 |

**Supplementary table 3. GBS cases during SARS-CoV-2 lockdown period (March-May) by year and region.**
